# PANACEA: a framework to maximise genetic diversity in genome-wide association study meta-analyses

**DOI:** 10.64898/2026.08.06.26359891

**Authors:** Chuan Fu Yap, Andrew P Morris

## Abstract

There have been recent efforts by the human genetics research community to increase the genetic diversity of participants contributing to genome-wide association studies (GWAS) of complex human traits and diseases. The traditional multi-ancestry GWAS approach is to first assign participants to continental ancestry labels based on their genetic similarity to individuals in reference datasets. Ancestry-specific GWAS are then conducted separately for each continental label, the results of which are aggregated through multi-ancestry meta-analysis. However, with this approach, a participant may be assigned to an ancestry group that does not reflect their personal view of ethnicity/race or may be excluded because their genetic ancestry is not sufficiently similar to individuals in reference datasets to be assigned to a single group. Here, we present a novel pipeline (PANACEA) for fully inclusive multi-ancestry meta-analysis that employs a continuous and multi-dimensional representation of ancestry that maximises the genetic diversity of GWAS. Through application to multi-ancestry GWAS of type 2 diabetes susceptibility and simulations, we demonstrate that the inclusive pooled analysis provides equivalent protection against population structure to a traditional ancestry-stratified analysis but, importantly, offers increased power to detect association through increased sample size by not excluding participants with outlying ancestry. The pooled inclusive analysis also enables assessment of ancestry-correlated heterogeneity in allelic effects without the need to assign participants to continental labels that may not sufficiently reflect genetic diversity within ancestry groups.

## INTRODUCTION

Genome-wide association studies (GWAS) have identified thousands of loci contributing to a wide range of complex human diseases^1,2^. However, most participants in these GWAS have genetic ancestry that is most similar to European ancestry individuals in the 1000 Genomes Project and/or the Human Genetic Diversity Project (1KGP/HGDP)^3-5^, which is in sharp contrast to the global burden of many diseases^6^. European ancestry individuals do not encompass the global diversity of genetic variation, which therefore limits the opportunities for clinical translation of GWAS (including drug target discovery and polygenic prediction) that will be beneficial for all individuals and therefore exacerbates health inequities^7,8^.

To address this bias, there have been recent efforts by the human genetics research community to increase the genetic diversity of participants contributing to GWAS, analyses of which require advanced multi-ancestry methods^9^. The traditional approach is to first assign participants to an ancestry group, typically represented by a continental label, based on their genetic similarity to individuals in 1KGP/HGDP. Ancestry-specific GWAS are then conducted within each ancestry group, the results of which are aggregated through multi-ancestry meta-analysis^10^. This approach expands the genetic diversity of GWAS participants and enables modelling of the heterogeneity in allelic effects between ancestry groups that could arise due to differences in patterns of linkage disequilibrium (LD), polygenic background, and exposure to environmental risk factors (if not accounted for in the analysis)^11^. However, with this approach, participants are assigned to distinct ancestry groups with labels that may not reflect personal views of ethnicity/race or fully represent the scale of genetic diversity within the group^12^. Furthermore, some admixed participants are excluded because their genetic ancestry is not sufficiently similar to individuals in 1KGP/HGDP to be assigned to a single group^13^.

In this study, we present a novel pipeline for fully inclusive multi-ancestry meta-analysis that employs a continuous and multi-dimensional representation of ancestry that maximises the genetic diversity of GWAS and removes the need for labels that may not be consistent with ancestry from social, historical, and cultural perspectives^14^. We investigate the properties of the novel pipeline through simulations and highlight the benefits over traditional ancestry-stratified analysis through application to multi-ancestry GWAS of type 2 diabetes (T2D) susceptibility.

## RESULTS

### Overview of methods

An overview of the novel pipeline for inclusive pooled analyses is presented in **Supplementary Figure 1**. To represent global human genetic diversity, we first derive 20 reference axes of genetic variation (AGVs) from principal components analysis (PCA) of whole-genome sequence obtained from 4,150 individuals from 1KGP/HGDP^5^ (**Figure 1, Supplementary Figure 2**). The first three reference AGVs capture genetic diversity across seven major ancestry groups defined by 1KGP/HGDP (African, American, Central and South Asian, East Asian, European, Middle Eastern, and Oceanian), whilst the remaining AGVs describe finer-scale genetic differences within these major ancestry groups. Loadings from the PCA are then used to project participants from each GWAS onto the reference AGVs. For each GWAS, pooled association analyses including all participants are next conducted using computationally efficient whole genome regression^15^, implemented in REGENIE, with adjustment for the projected reference AGVs to control for population structure.

**Figure 1.**
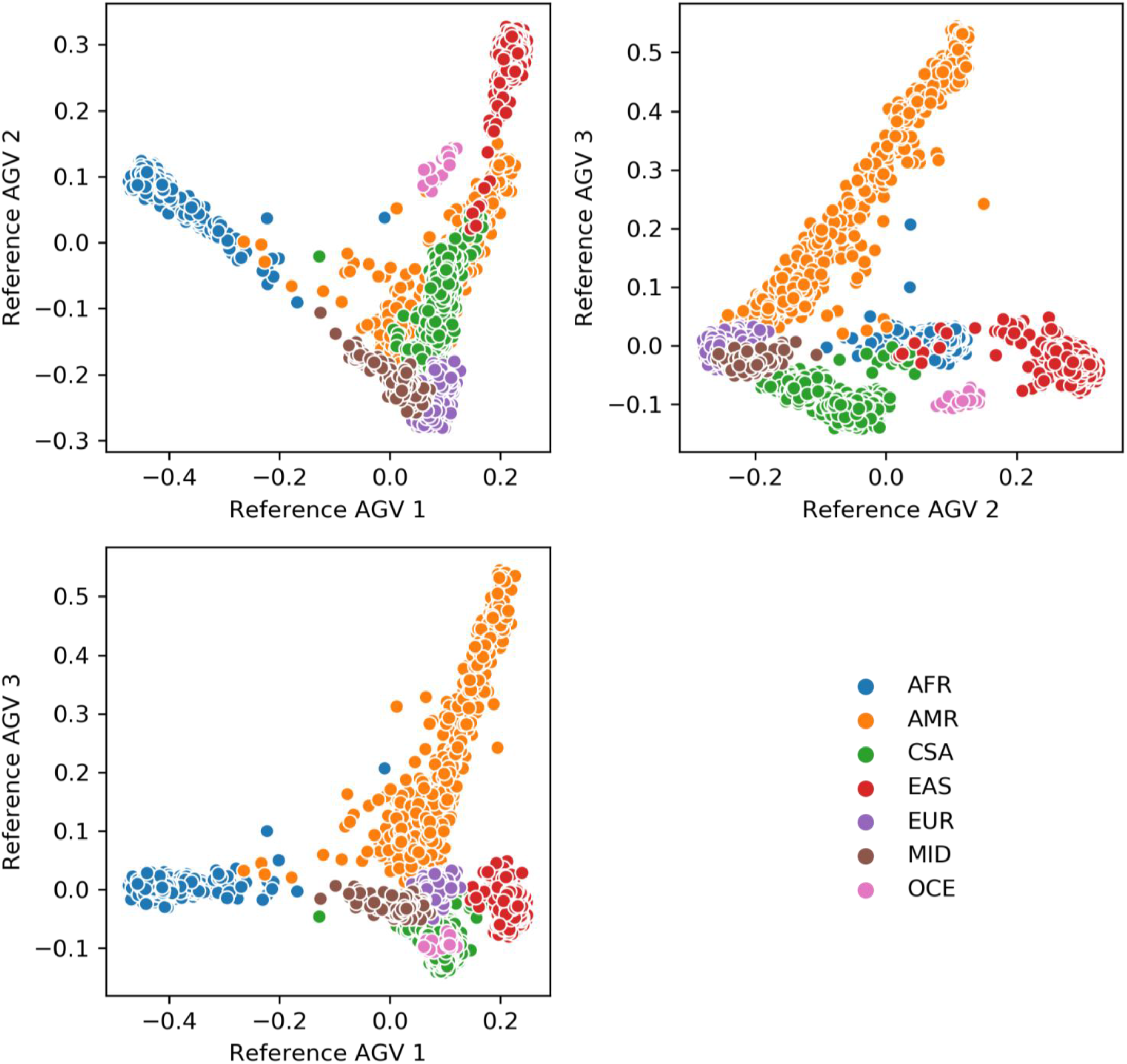
Reference axes of genetic variation (AGVs) derived from 1KGP/HGDP reference data. In each panel, points correspond to individuals from 1KGP/HGDP, plotted according to their position on the first three reference AGVs, obtained from principal components analysis of LD-pruned autosomal variants from whole-genome sequence data. Each individual is coloured according to self-reported continental ancestry group: African (AFR), American (AMR), Central and South Asian (CSA), East Asian (EAS), European (EUR), Middle Eastern (MID), and Oceanian (OCE).

Heterogeneity in allelic effects that is correlated with ancestry is then modelled within each GWAS by including an interaction with the first three reference AGVs using output from REGENIE. Association summary statistics from each GWAS can be aggregated via meta-analysis under an inverse-variance weighted fixed-effects model for the allelic effect^16^ and via synthesis of regression slopes for heterogeneity effect estimates^17^. The methodology has been implemented in PANACEA (Pooled ANAlysis with a Continuous Exposition of Ancestry) available at: https://github.com/chuanfuyap/PANACEA. Full details of the approach are provided in **Methods**.

### GWAS of T2D susceptibility

To demonstrate the utility of the novel pipeline for inclusive pooled analyses, and highlight advantages over traditional “ancestry-stratified” methods, we considered GWAS of T2D susceptibility in two population biobanks that include participants of diverse genetic ancestry (**Supplementary Table 1**): the UK Biobank (UKB)^18^ and the Resource for Genetic Epidemiology Research in Adult Health and Aging (GERA)^19^.

Participants from UKB were genotyped with one of two genotyping arrays, jointly imputed to a combined reference panel from the 1000 Genomes Project, UK10K Project, and Haplotype Reference Consortium^20^. After quality control, UKB comprised phenotype and genotype data in 486,989 participants at 10,936,942 variants with minor allele frequency (MAF) ≥0.5% and imputation quality (info) ≥0.8. Participants from GERA were genotyped with one of four ancestry-aware genotyping arrays (European, African, Latino, East Asian), with each array separately imputed to the Trans-Omics for Precision Medicine (TOPMed) reference panel^21^. After quality control, GERA comprised a total of 71,604 participants at up to 16,033,228 variants (variable by genotyping array) with MAF ≥0.5% and imputation quality (*r*^2^) ≥0.8.

We projected all participants from UKB and GERA onto the 20 reference AGVs (**Supplementary Figure 3**). Within each GWAS (one for UKB and four for GERA, stratified by genotyping array), we used all participants to conduct a pooled test of association of each variant with T2D status using whole genome regression implemented in REGENIE with adjustment for sex and the projected reference AGVs as covariates. For comparison, we implemented a traditional ancestry-stratified analysis for which participants were first assigned to continental ancestry groups (African, American, Central and South Asian, East Asian, European) within each GWAS based on self-reported ethnicity and GWAS-specific AGVs (**Methods, Supplementary Figure 4, Supplementary Table 2**). A total of 19,395 (4.0%) participants in the UKB GWAS and 5,415 (7.6%) participants across the four array-stratified GERA GWAS could not be assigned to an ancestry group and were excluded from the traditional ancestry-stratified analysis. Within each GWAS, we conducted ancestry-stratified tests of association of each variant with T2D status using whole genome regression implemented in REGENIE with adjustment for sex. For both the pooled and ancestry-stratified analyses, allelic effects for each variant were then aggregated across GWAS using inverse-variance weighted fixed-effects meta-analysis (**Supplementary Figure 5**).

We observed no difference in genome-wide inflation in association summary statistics between the pooled and ancestry-stratified analysis approaches, with genomic control inflation factors of λ=1.159 for both (**Supplementary Figure 6**). These results suggest that adjustment for reference AGVs in the inclusive pooled analysis provides equivalent control for population structure as GWAS-specific AGVs in the traditional ancestry-stratified analysis, but has the advantage of including all participants, irrespective of ancestry.

We identified a total of 158 and 146 lead variants at genome-wide significance (*P*<5×10^-8^) in the inclusive pooled and traditional ancestry-stratified analyses, respectively (**Supplementary Tables 3 and 4**). For both analysis approaches, the strongest signals of association mapped to well-established loci for T2D susceptibility^22^, including *TCF7L2, CDKAL1, IGF2BP2, SLC30A8, FTO, KCNQ1*, and the genomic regions including *CDKN2A* and *CDKN2B*, and *HHEX* and *IDE*. Of the 96 lead variants reported for both methods, 66 (68.8%) demonstrated stronger signals of association in the inclusive pooled analysis than the traditional ancestry-stratified analysis (**Figure 2**). For example, stronger signals of association were observed in the inclusive pooled analysis for the lead variants mapping to *CDKAL1* (rs9368222, pooled analysis *P*=2.0×10^-59^, ancestry-stratified analysis *P*=2.3×10^-54^), the region including *HHEX* and *IDE* (rs7898054, pooled analysis *P*=2.1×10^-46^, ancestry-stratified analysis *P*=2.0×10^-41^), *SLC30A8* (rs3802177, pooled analysis *P*=3.1×10^-42^, ancestry-stratified analysis *P*=4.1×10^-40^), and *FTO* (rs56094641, pooled analysis *P*=1.8×10^-41^, ancestry-stratified analysis *P*=1.1×10^-47^).

**Figure 2.**
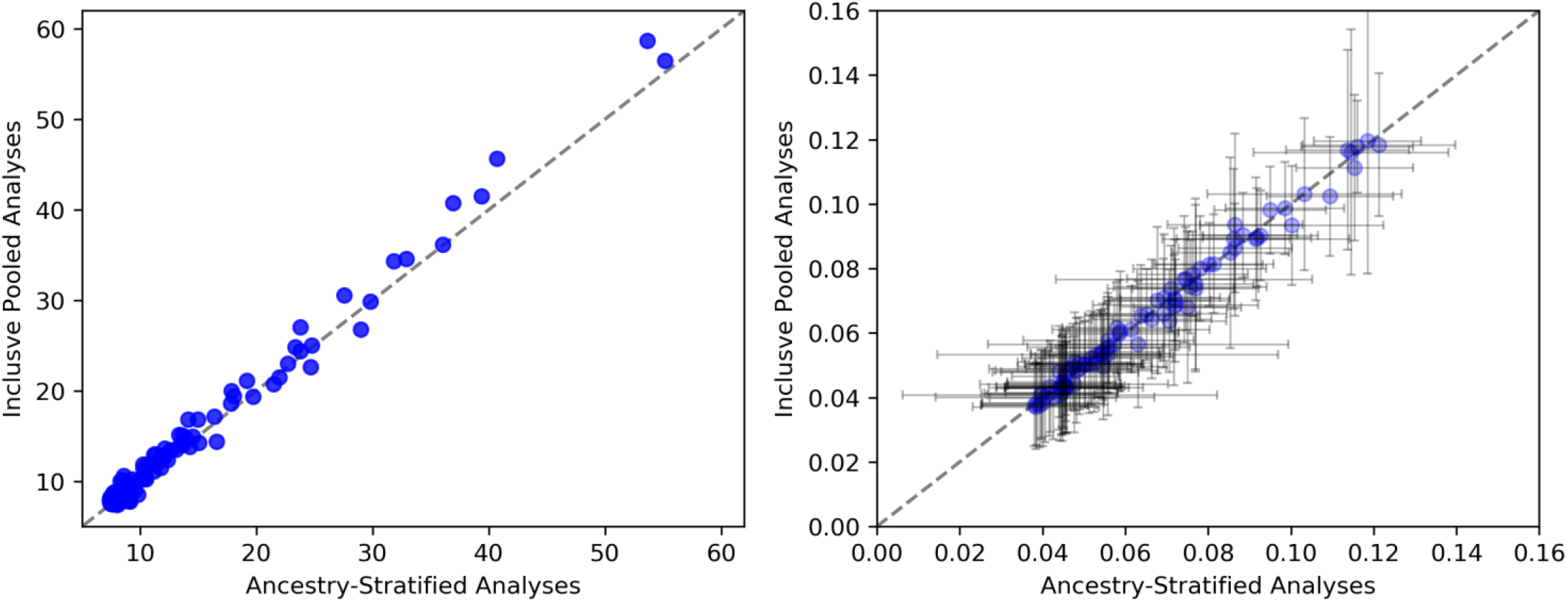
Comparison of association summary statistics for T2D for 96 lead variants identified in inclusive pooled analysis and ancestry-stratified analysis in the UK Biobank and the Resource for Genetic Epidemiology Research in Adult Health and Aging. In each panel, points correspond to lead variants, plotted according to association summary statistics from the ancestry-stratified analysis on the x-axis and from the inclusive pooled analysis on the y-axis. (a) *P*-values on a -log_10_ scale. (b) Allelic log-odds ratio (for the T2D risk allele), where the grey lines represent 95% confidence intervals. Variants with log-odds ratio >0.2 (rs147581833 at the *ANKH* locus and rs76895963 at the *CCND2* locus) are excluded for ease of presentation.

We next tested for evidence of ancestry-correlated heterogeneity in allelic effects on T2D susceptibility at the 158 lead variants from the inclusive pooled analysis by testing for evidence of an interaction with the first three reference AGVs using the output from REGENIE, aggregating heterogeneity effect estimates across via synthesis of regression slopes. We identified 14 variants with significant evidence of ancestry-correlated heterogeneity (*P*<0.05) from the interaction test (**Supplementary Table 5**). Consistent with previous reports^22,23^, we observed significant enrichment of ancestry-correlated heterogeneity at lead variants for T2D susceptibility (expected 7.9 variants, binomial test *P*=0.028). The strongest evidence for ancestry-correlated heterogeneity was observed for the lead variant at the *TCF7L2* locus (rs35198068, *P*=1.0×10^-4^). The effect of the risk allele at this variant was least in participants from UKB and GERA who are genetically most similar to East Asian ancestry individuals in 1KGP/HGDP and greatest in those who are genetically most similar to European ancestry individuals (**Figure 3**). There was also a gradient of the effect of the risk allele in participants from UKB and GERA who are genetically most similar to American ancestry individuals in 1KGP/HGDP. The effect was least in those most genetically similar to Pima, Karitiana, Maya, and Surui individuals (**Supplementary Figure 7**), emphasizing the importance of a more continuous view of genetic ancestry than cannot be accommodated with continental ancestry labels.

**Figure 3.**
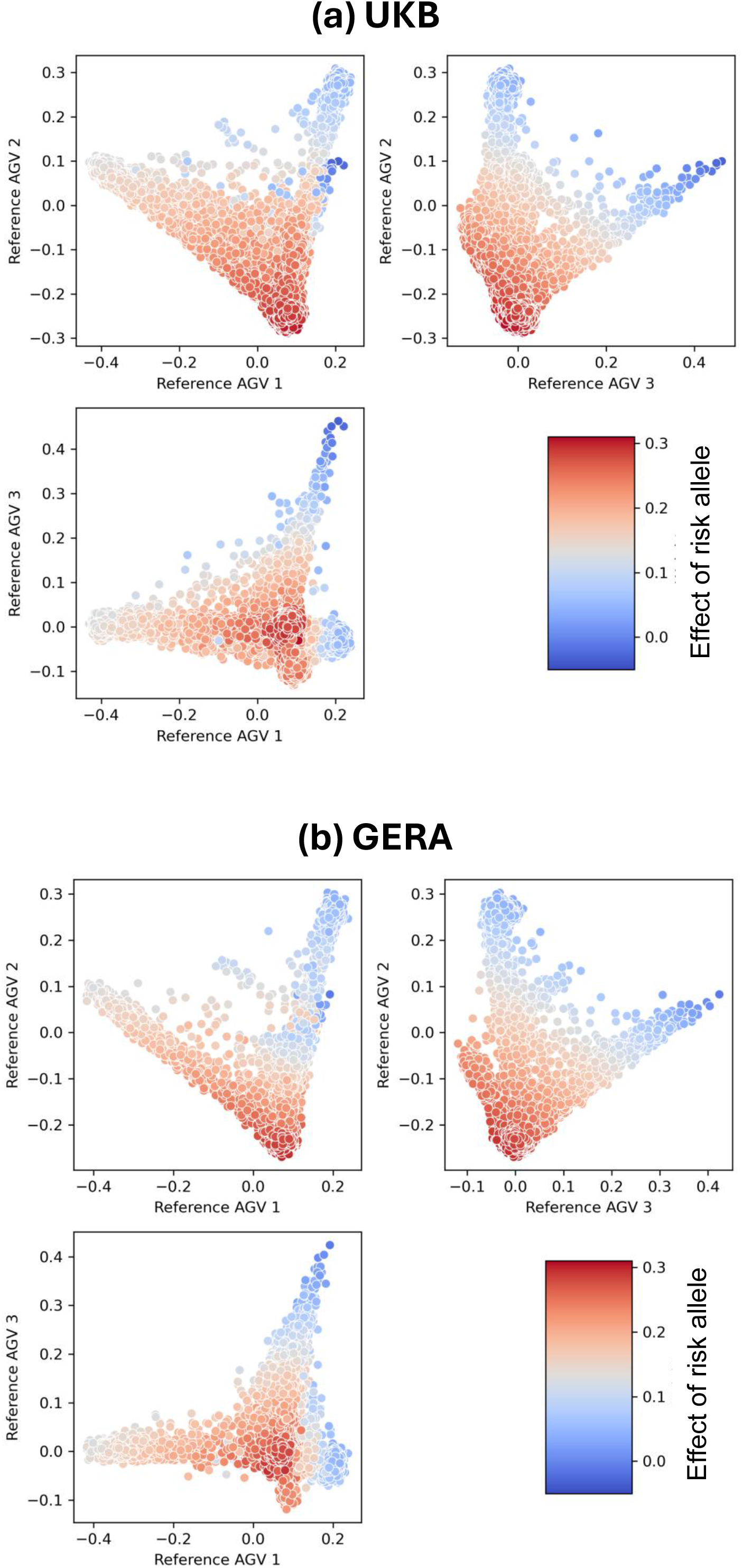
Ancestry-correlated heterogeneity in the effect of the risk allele on T2D susceptibility at the lead variant, rs35198068, at the *TCF7L2* locus. (a) Participants in the UK Biobank (UKB). (b) Participants in the Resource for Genetic Epidemiology Research in Adult Health and Aging (GERA). In each panel, each point corresponds to a participant, plotted according to their projection onto the first three axes of genetic variation (AGVs) derived from 1KGP/HGDP (described in Figure 2). Each point is coloured by the expected effect of the risk allele on T2D susceptibility, given the projection of the participant onto the first three reference AGVs.

### Simulation study

To demonstrate the relative performance of the inclusive pooled GWAS analysis and the traditional ancestry-stratified GWAS analysis, we conducted extensive simulations based on participants in UKB. For the inclusive pooled analysis, 486,989 participants were projected onto the 20 reference AGVs derived from 1KGP/HGDP. For the traditional ancestry-stratified GWAS, 467,594 participants were allocated to four ancestry groups (African, Central and South Asian, East Asian, European), with 19,395 participants excluded as outliers (**Methods**). We considered a quantitative trait determined by a causal variant and an unobserved covariate that varies with ancestry. The trait model was parameterised in terms of the marginal effects of the trait increasing allele and covariate, denoted *β*_*G*_ and *β*_*X*_, respectively, and the interaction between the trait increasing allele and covariate, denoted covariate, *β*_*GX*_. For each parameter combination, we generated 1,000 replicates of genotype and phenotype data, each based on the random selection of an imputed variant (MAF ≥0.5%, info ≥0.8) as causal. In each replicate of data, we used all participants to conduct: (i) a pooled test of association of the causal variant with the trait using whole genome regression implemented in REGENIE with adjustment for the projected reference AGVs as covariates; and (ii) a test of interaction of the causal variant with the first three reference AGVs using the output from REGENIE. We also conducted an ancestry-stratified test of association of the causal variant with the trait using whole-genome regression implemented in REGENIE, with allelic effects aggregated across ancestry groups using inverse-variance weighted fixed-effects meta-analysis.

We began by considering the type I error rates of the inclusive pooled test of association and ancestry-stratified test of association under the null model (i.e. *β*_*G*_ = *β*_*GX*_ = 0). At a nominal threshold of significance (*P*<0.05), we observed no inflation in type I error rates for either test (**Supplementary Table 6**). These results support the observation from our GWAS analyses of T2D susceptibility that the adjustment for reference AGVs in the whole genome regression model is sufficient to account for population structure in the inclusive pooled analysis, despite the increased genetic diversity of participants.

We next compared the power to detect association with the causal variant, at genome-wide significance (*P*<5×10^-8^), with the inclusive pooled analysis and ancestry-stratified analysis (**Figure 4**). In the absence of an interaction between the causal variant and ancestry-correlated covariate (*β*_*GZ*_ = 0), we observed an increase in power for the inclusive pooled analysis over the ancestry-stratified analysis, irrespective of the effect size of the ancestry-correlated covariate. This improvement in power would be expected for the increased sample size offered by not excluding participants who could not be allocated to an ancestry group. The increased power for the inclusive pooled analysis over the ancestry-stratified analysis was also observed in the presence of an interaction between the causal variant and ancestry-correlated covariate (*β*_*GZ*_ = 0.1, **Supplementary Figure 8**).

**Figure 4.**
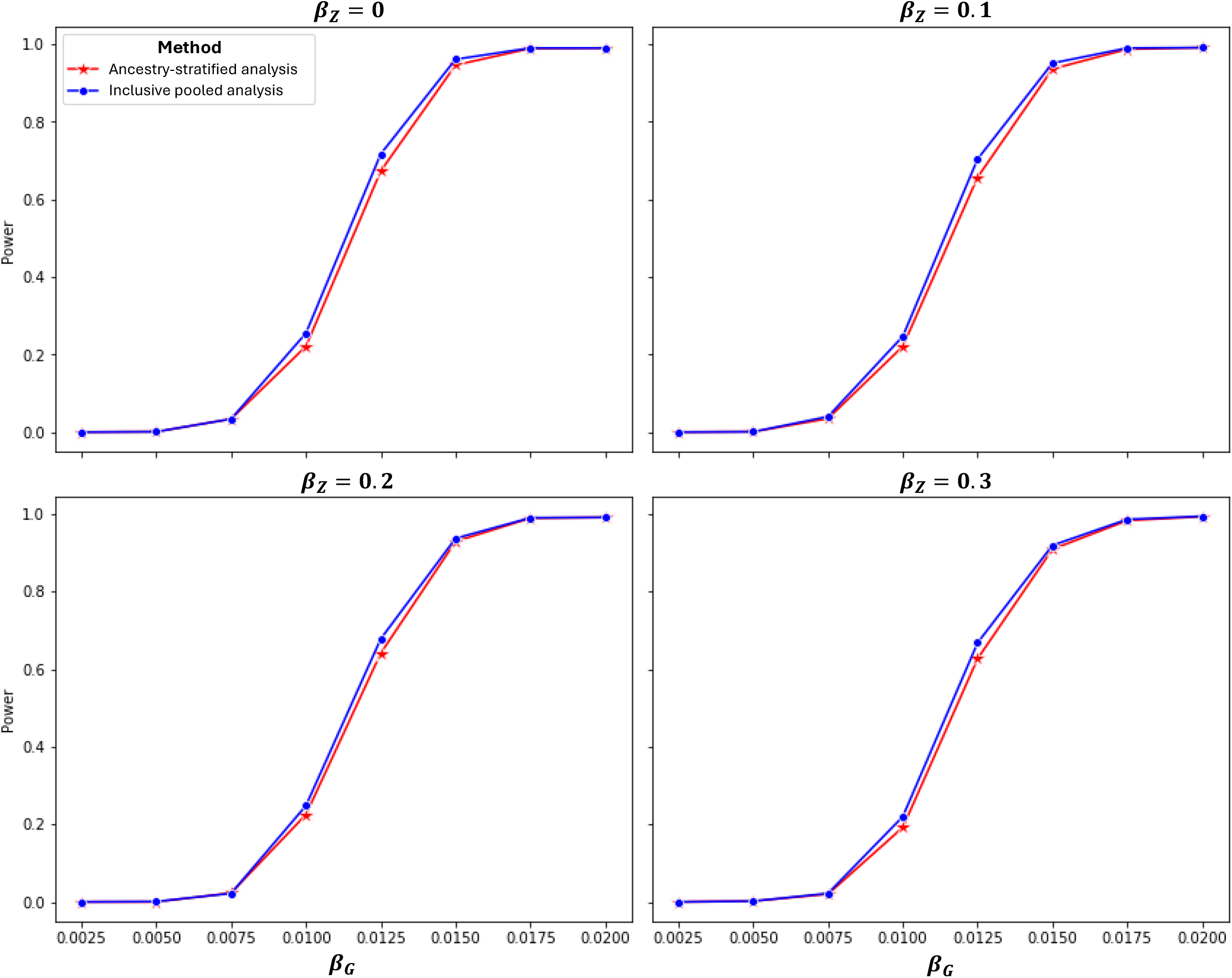
Power to detect association (at genome-wide significance, *P*<5×10^-8^) with causal variants for inclusive pooled analysis and ancestry-stratified analysis based on participants in the UK Biobank. Power is presented for each method as a function of the marginal effect of the causal variant, *β*_*G*_, for four different marginal effects of an unobserved ancestry-correlated covariate, *β*_*Z*_ = {0, 0.1, 0.2, 0.3}.

Finally, we assessed the power to detect ancestry-correlated heterogeneity in allelic effects with the inclusive pooled analysis in the presence of an interaction between the causal variant and the ancestry-correlated covariate. For a fixed marginal effect of the ancestry-correlated covariate, the power to detect ancestry-correlated heterogeneity at a nominal threshold of significance (*P*<0.05) increased with the interaction effect of the causal variant with the covariate, irrespective of the marginal effect of the causal variant (**Supplementary Figure 9**). These results highlight the power of the approach to detect heterogeneity in allelic effects driven by interaction of a causal variant with an unobserved covariate that varies with ancestry.

## DISCUSSION

We have developed a novel inclusive pipeline (PANACEA) for multi-ancestry GWAS (meta-)analysis that overcomes the challenge of assigning participants to continental ancestry labels by considering a continuous and multi-dimensional representation of genetic ancestry, which better reflects the continuum of human genetic diversity and demographic history^14^. Within this framework, we increase power to detect association by not excluding participants that cannot be assigned to a continental label, while controlling for population structure that could lead to an increase in type I error rates. We also allow for assessment of ancestry-correlated heterogeneity in allelic effects without the need to assign participants to continental labels that may not sufficiently reflect genetic diversity within ancestry groups.

Recent investigations have demonstrated that the inclusive pooled approach is a powerful and scalable strategy for multi-ancestry GWAS, increasing power for genetic discovery while maintaining rigorous population structure control^24^. The novelty of our pipeline is the projection of participants onto reference AGVs, which has the advantage of avoiding bias against under-represented ancestry groups within the GWAS^25^ and harmonising heterogeneity effect estimates and adjustments for population structure across GWAS to enable meta-analysis. The modelling of ancestry-correlated heterogeneity as a function of AGVs has been previously utilised in the context of multi-ancestry GWAS meta-regression, implemented in the MR-MEGA software, and has been shown to increase power to detect association over fixed-effects meta-analysis^11^.

The PANACEA pipeline has been developed for use with whole-genome regression, implemented in REGENIE, which in addition to adjustment for reference AGVs as covariates, offers correction for relatedness and population structure. In principle, other GWAS analysis software that accounts for relatedness between participants can be used in place of REGENIE, such as SAIGE^26^. However, REGENIE is computationally efficient, can be applied to both quantitative and binary phenotypes, and can allow for case-control imbalance. Whilst REGENIE cannot be used to fit interaction models for variants with multiple covariates, simultaneously, output can be directly used to test for interaction with the first three reference AGVs to assess the evidence for ancestry-correlated heterogeneity within the PANACEA pipeline.

An advantage of modelling ancestry-correlated in the PANACEA pipeline is that we can estimate the allelic effect on a trait/disease at a variant for an individual that is projected onto AGVs. Such allelic effect estimates would allow for the genetic ancestry of the individual and could be used as weights in polygenic scores in place of weights that are derived from fixed-effects multi-ancestry meta-analysis or ancestry-specific meta-analysis that are typically estimated at the level of continental group. Our application of the inclusive pooled analysis approach to GWAS of T2D susceptibility in UKB and GERA highlighted strong evidence of ancestry-correlated heterogeneity in allelic effects for the lead variant at the *TCF7L2* locus. Amongst participants who are genetically most similar to American ancestry individuals in 1KGP/HGDP, there was a gradient of allelic effects that would not be captured using the continental ancestry label. Modelling heterogeneity as a function of reference AGVs could, therefore, provide more accurate weights and improved performance of polygenic scores across the spectrum of genetic diversity^27,28^.

## Data Availability

All data produced in the present study are available upon reasonable request to the authors

https://github.com/chuanfuyap/PANACEA

## RESOURCE AVAILABILITY

### Lead contact

Requests for further information and resources should be directed to and will be fulfilled by the lead contact, Andrew Morris.

### Materials availability

This study did not generate any new materials.

### Data and code availability

The 1KGP/HGDP reference data can be download from https://gnomad.broadinstitute.org/data#v3-hgdp-1kg. GERA data can be accessed through dbGaP (https://dbgap.ncbi.nlm.nih.gov/home, accession number phs000674.v1.p1). UBK data can be accessed through Access to UK Biobank can obtained through https://www.ukbiobank.ac.uk/use-our-data/apply-for-access/. Reference AGV loadings and analysis scripts used in this manuscript are available from the PANACEA github at https://github.com/chuanfuyap/PANACEA.

## ACNOWLEDGEMENTS

This work was supported by Versus Arthritis (21754), NIHR Manchester Biomedical Research Centre (NIHR203308) and UKRI MRC (MR/W029626/1). The authors acknowledge the assistance of Research IT Support and the use of the Computational Shared Facility at The University of Manchester. We thank Dr Joelle Mbatchou for discussion on the implementation of the ancestry-correlated heterogeneity model in REGENIE.

This research has been conducted using data from UK Biobank, a major biomedical database under project application 117222. GERA data came from a grant, the Resource for Genetic Epidemiology Research in Adult Health and Aging (RC2 AG033067; Schaefer and Risch, PIs) awarded to the Kaiser Permanente Research Program on Genes, Environment, and Health (RPGEH) and the UCSF Institute for Human Genetics. The RPGEH was supported by grants from the Robert Wood Johnson Foundation, the Wayne and Gladys Valley Foundation, the Ellison Medical Foundation, Kaiser Permanente Northern California, and the Kaiser Permanente National and Northern California Community Benefit Programs. The RPGEH and the Resource for Genetic Epidemiology Research in Adult Health and Aging are described in the following publication, Schaefer C, et al., The Kaiser Permanente Research Program on Genes, Environment and Health: Development of a Research Resource in a Multi-Ethnic Health Plan with Electronic Medical Records, In preparation, 2013.

## AUTHOR CONTRIBUTIONS

APM conceived and supervised the study and developed the methodological concepts. CFY implemented the methodology and conducted data analysis and simulations. APM and CFY wrote the manuscript.

## METHODS

### Derivation of reference axes of genetic variation

Reference axes of genetic variation were derived from whole-genome sequence data of 4,150 individuals from 1KGP/HGDP (downloaded from https://gnomad.broadinstitute.org/data#v3-hgdp-1kg, NCBI hg38) that represent genetic diversity across seven major ancestry groups^5^. The following quality control steps were performed: exclusion of related individuals not in the maximal independent set (kinship statistic of 0.05) using PC Relate^29^ implemented in Hail (https://hail.is/docs/0.2/methods/relatedness.html#hail.methods.pc_relate, https://hail.is/docs/0.2/methods/misc.html#hail.methods.maximal_independent_set); retention of autosomal variants with MAF ≥5%; exclusion of variants that are not present in 1KGP. Known long-range LD regions were excluded, and variants were LD-pruned (*r*^2^<0.01) using PLINKv1.9^30^. PCA was applied to the LD-pruned variants using Hail (https://hail.is/docs/0.2/methods/stats.html#hail.methods.pca). Loadings from the first 20 AGVs were retained for downstream analysis and are available at: https://github.com/chuanfuyap/PANACEA/tree/main/gnomad_loadings.ht.

### Inclusive pooled GWAS (meta-)analysis approach

Scripts for analysis are available at: https://github.com/chuanfuyap/PANACEA. Variants that overlap between the LD-pruned set for which loadings are reported and those passing quality control in the GWAS are extracted, and alleles aligned to the reference, using https://github.com/chuanfuyap/PANACEA/blob/main/extract_fix_ref.sh. The loadings are then applied to genotypes at the overlapping variants to project GWAS participants onto the first 20 AGVs using https://github.com/chuanfuyap/PANACEA/blob/main/pca_projection.sh.

We next test for association of each variant with the phenotype using whole-genome regression^15^, implemented in REGENIE (https://rgcgithub.github.io/regenie/), with additive genotype dosage and adjustment for covariates and the first 20 projected AGVs. For each variant, REGENIE fits the model

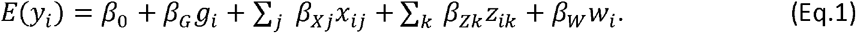

In this equation, for the *i*th individual, *g*_*i*_ is the additive genotype dosage of the coded allele, *x*_*i*_ denotes values of covariates, *z*_*i*_ denotes the projection onto reference AGVs, and *w*_*i*_ is the genetic prediction from a “leave one chromosome out” approach. REGENIE allows for analysis of quantitative traits under a linear regression model and binary phenotypes under a logistic regression (including the option of a Firth correction to allow for case-control imbalance).

To test for evidence of ancestry-correlated heterogeneity of the allelic effect at a variant, we extend (Eq.1) to include an interaction of genotype with projections onto the first three AGVs, given by

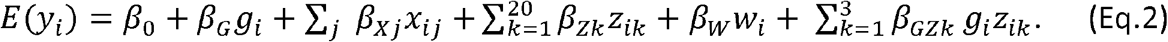

REGENIE currently allows for testing a genotype interaction with only a single covariate. We have therefore implemented a likelihood ratio test, https://github.com/chuanfuyap/PANACEA/blob/main/regintest.sh, by comparing the two models, which makes use of the genetic predictions from REGENIE.

For each variant, we aggregate allelic effect estimates, *β*_*G*_, across GWAS under an inverse-variance fixed-effects model implemented in METAL^16^ (https://genome.sph.umich.edu/wiki/METAL_Program). We also aggregate heterogeneity effect estimates, *β*_*GZ*_, across GWAS via synthesis of regression slopes^17^, implemented in https://github.com/chuanfuyap/PANACEA/blob/main/multivar-meta.sh. Finally, we estimate the effect of the coded allele for the *i*th individual, *b*_*i*_, as a function of their projection onto the first three reference AGVs, given by

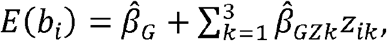

where 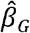 is the marginal allelic effect estimate and 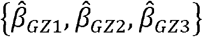 are heterogeneity effect estimates from the synthesis of regression slopes.

### GWAS of T2D susceptibility in UK Biobank

UKB is a detailed prospective study of more than 500,000 participants that were aged 40-69 years when recruited between 2006 and 2010^18^. Extensive data have been collected from participants, including health and lifestyle questionnaires, physical measurements, and follow-up for a wide range of health-related outcomes, together with samples for DNA extraction. Genome-wide genotyping was undertaken using the Affymetrix UKB or BiLEVE arrays, with genotype calling and quality control conducted by the UKB Analysis Team^20^. Subsequent imputation was performed up to reference panels from the 1000 Genomes Project^3^, UK10K Project^31^, and Haplotype Reference Consortium^32^. Data were accessed under project application 117222. We retained variants with MAF ≥0.5% and imputation quality (info) ≥0.8 for downstream association analysis. T2D status was derived from data-field 130708, with a participant defined as a case if they had a recorded date of first occurrence of non-insulin-dependent diabetes mellitus, and all non-cases defined as controls.

### GWAS of T2D susceptibility in GERA

GERA is a large multi-ethnic population-based cohort of more than 100,000 adults who are members of the Kaiser Permanente Medical Care Plan, Northern California Region, and participants in its Research Program on Genes, Environment, and Health. Data were accessed from dbGaP via accession phs000674.p1. GERA participants have been genotyped using one of four custom arrays, which have been designed to maximise coverage of common and low-frequency variants in European, East Asian, African, and Latino individuals^33,34^. Quality control of the genotype data was conducted within each array separately and has been previously described^19^. Briefly, we removed individuals from known pedigrees and/or with call rate <97%, and excluded variants with call rate <95% and extreme deviation from Hardy-Weinberg equilibrium (autosomes only, exact *P*<10^-6^). Each array was then separately imputed to the TOPMed r3 reference panel using the TOPMed Imputation Server^21,35^ (https://imputation.biodatacatalyst.nhlbi.nih.gov/). We retained variants with MAF ≥0.5% and imputation quality (*r*^2^) ≥0.8 for downstream association analysis. T2D status was derived from summarising ICD-9 coded diagnoses in Kaiser Permanente’s electronic medical records, with a participant defined as a case by the occurrence of two or more T2D diagnoses occurring on separate days, and all non-cases defined as controls.

### Traditional ancestry-stratified GWAS analysis of T2D susceptibility across UKB and GERA

UKB and GERA participants were first allocated to ancestry groups. In UKB, we utilised genetic principal components provided in data-field 22009 and top-level self-reported ethnicity reported in data-field 21000 (Asian, Black, Chinese, White). In GERA, we merged imputed variants across the four arrays and generated genetic principal components by applying PCA to LD-pruned autosomal variants (MAF ≥5%, *r*^2^<0.01) using PLINKv1.9^30^, and used self-reported ethnicity (Asian, Black, Hispanic, White). For each self-reported ethnic group, separately for UKB and GERA, a multivariate Normal distribution was generated across the first three genetic principal components. The Mahalanobis distance was then calculated for each participant relative to the group-specific distribution. Participants with distance less than two standard deviations from the group mean along each genetic principal component were allocated to the group, with all other participants excluded as outliers. Association analyses were then conducted separately for UKB and for each array in GERA. Within each allocated ancestry group, we tested for association of each variant with T2D using whole-genome regression^15^, implemented in REGENIE, under a Firth-corrected logistic regression model with additive genotype dosage and adjustment for sex. For each variant, association summary statistics from each ancestry-stratified GWAS in UKB and each array in GERA were aggregated via meta-analysis under an inverse-variance weighted fixed-effects model for the marginal allelic effect, implemented in METAL^16^. Lead variants attaining genome-wide significance (*P*<5×10^-8^) were identified by distance-based clumping (2Mb window) and mapped to loci reported in the largest published multi-ancestry GWAS meta-analysis of T2D susceptibility^22^.

### Inclusive pooled GWAS analysis of T2D susceptibility across UKB and GERA

Association analyses were conducted separately for UKB and for each array in GERA. Each participant was projected onto the first 20 reference AGVs using https://github.com/chuanfuyap/PANACEA/blob/main/pca_projection.sh. We tested for association of each variant with T2D using whole-genome regression^15^, implemented in REGENIE, under a Firth-corrected logistic regression model with additive genotype dosage and adjustment for sex and the first 20 projected reference AGVs. For each variant, we also tested for heterogeneity in allelic effects on T2D that is correlated with ancestry by testing for interaction with the first three projected reference AGVs using https://github.com/chuanfuyap/PANACEA/blob/main/regintest.sh. For each variant, association summary statistics from GWAS in UKB and each array in GERA were aggregated via meta-analysis under an inverse-variance weighted fixed-effects model for the marginal allelic effect, implemented in METAL^16^. For each variant, association summary statistics from GWAS in UKB and each array in GERA were aggregated using synthesis of regression slopes for heterogeneity effect estimates^17^, implemented in https://github.com/chuanfuyap/PANACEA/blob/main/multivar-meta.sh. Lead variants attaining genome-wide significance (*P*<5×10^-8^) were identified by distance-based clumping (2Mb window) and mapped to loci reported in the largest published multi-ancestry GWAS meta-analysis of T2D susceptibility^22^.

### Simulation study

To demonstrate the relative performance of the inclusive pooled GWAS analysis and the traditional ancestry-stratified GWAS analysis, we conducted extensive simulations based on participants in UKB. We considered the projection of each participant onto the reference AGVs and their allocation to an ancestry group (African, Central and South Asian, East Asian, European) as described above. We assumed a quantitative trait, *Y*, dependent on genotypes at a causal variant, *G*, coded under an additive genetic model, an unobserved covariate, *z*, and an interaction between *G* and *z*. Specifically, for the *i*th participant,

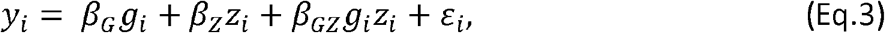

where *β*_*G*_ is the marginal effect of the causal variant, *β*_*X*_ is the marginal effect of the covariate, *β*_*G X*_ is the interaction effect between the causal variant and covariate, and *ε*_*i*_ ∼*N* 0,1). We assumed that the covariate varied with ancestry, which induced ancestry-correlated heterogeneity in the presence of a non-zero interaction, *β*_*GZ* ≠_ 0 between *G* and *Z*. Specifically, for the *i*th participant,

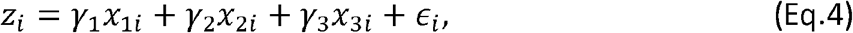

where *x*_*ji*_ is their projection onto the *j*th reference AGV, as defined in the GWAS of T2D susceptibility, *γ*_*j*_ is the effect of the *j*th reference AGV on the covariate, and *ϵ*_*i*_ ∼ *N* 0,1). Throughout, we assumed that *γ*_*l*_ = *γ*= *γ*= 0.1.

For each combination of simulation parameter values, *β*_*G*_, *β*_*Z*_, *β*_*GZ*_, we generated 1,000 replicates of data for all UKB participants. For each replicate, we first randomly selected a variant with MAF ≥0.5% and imputation quality (info) ≥0.8 as causal. We then simulated values of the covariate for each participant (Eq.4) and next simulated their phenotype, conditional on their covariate value and genotype at the causal variant (Eq.3). For the inclusive pooled GWAS analysis, we tested for association of the causal variant using whole-genome regression^15^, implemented in REGENIE, under a linear regression model with additive genotype dosage and adjustment for the first 20 projected reference AGVs. We also tested for heterogeneity in allelic effects that is correlated with ancestry by testing for interaction with the first three projected reference AGVs using https://github.com/chuanfuyap/PANACEA/blob/main/regintest.sh. For the traditional ancestry-stratified GWAS analysis, we tested for association of the causal variant using whole-genome regression^15^, implemented in REGENIE, under a linear regression model with additive genotype dosage, separately for each ancestry group, using the allocations defined in the GWAS of T2D susceptibility. Allelic effect estimates for the causal variant from each ancestry-stratified GWAS were finally aggregated via meta-analysis under an inverse-variance weighted fixed-effects model, implemented in METAL^16^.

