## Supplemental Figures for "PANACEA: a framework to maximise genetic diversity in genome-wide association study meta-analyses"

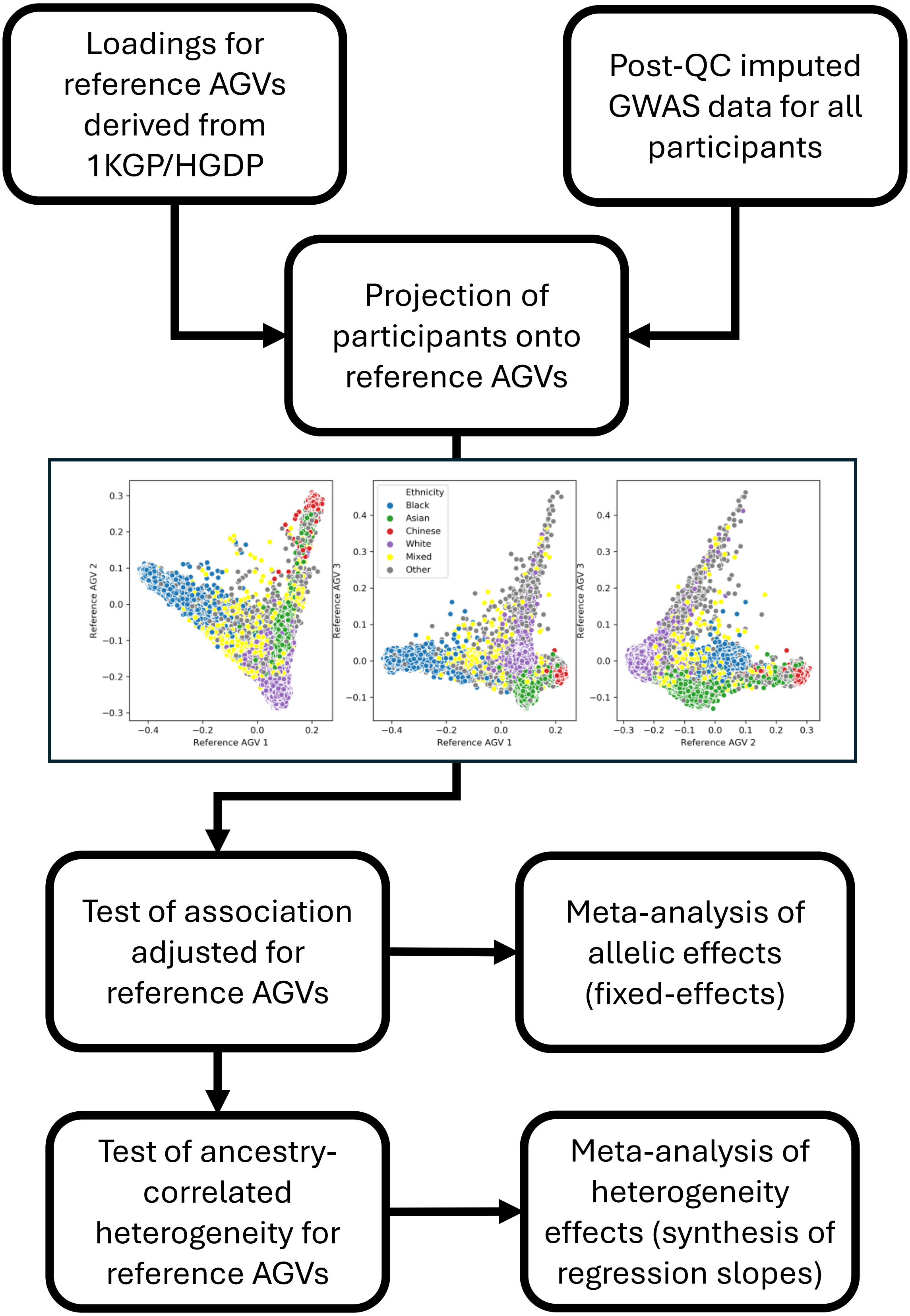


**Supplementary Figure 1. Inclusive pooled GWAS analysis implemented in the PANACEA pipeline.** Loadings derived from 1KGP/HGDP are used to project participants from a GWAS onto reference axes of genetic variation (AGVs). Pooled association analysis including all participants is conducted with adjustment for the projected reference AGVs to control for population structure. Heterogeneity in allelic effects that is correlated with ancestry is then modelled by including an interaction with the first three reference AGVs. Association summary statistics from multiple GWAS can be aggregated via meta-analysis under an inverse-variance weighted fixed-effects model for allelic effect estimates and via synthesis of regression slopes for heterogeneity effect estimates.

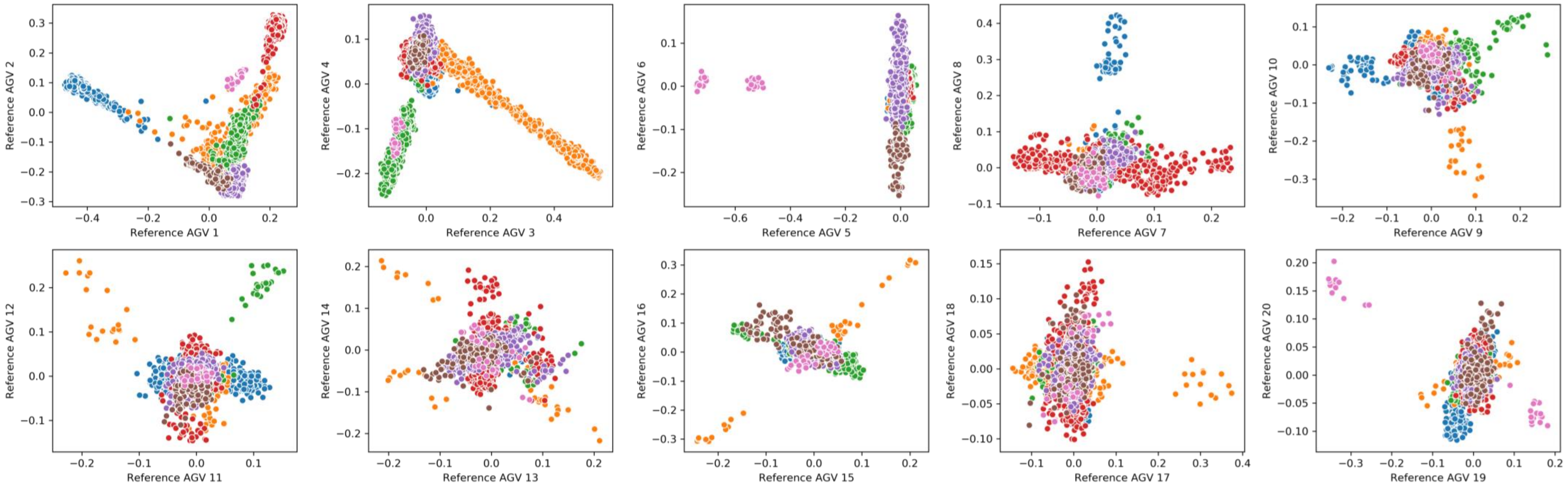


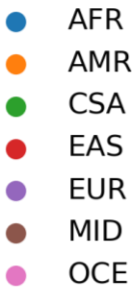


**Supplementary Figure 2. Reference axes of genetic variation (AGVs) derived from 1KGP/HGDP reference data.** In each panel, points correspond to individuals from 1KGP/HGDP, plotted according to their position on the first 20 AGVs, obtained from principal components analysis of LD-pruned autosomal variants from whole-genome sequence data. Each individual is coloured according to self-reported continental ancestry group: African (AFR), American (AMR), Central and South Asian (CSA), East Asian (EAS), European (EUR), Middle Eastern (MID), and Oceanian (OCE).

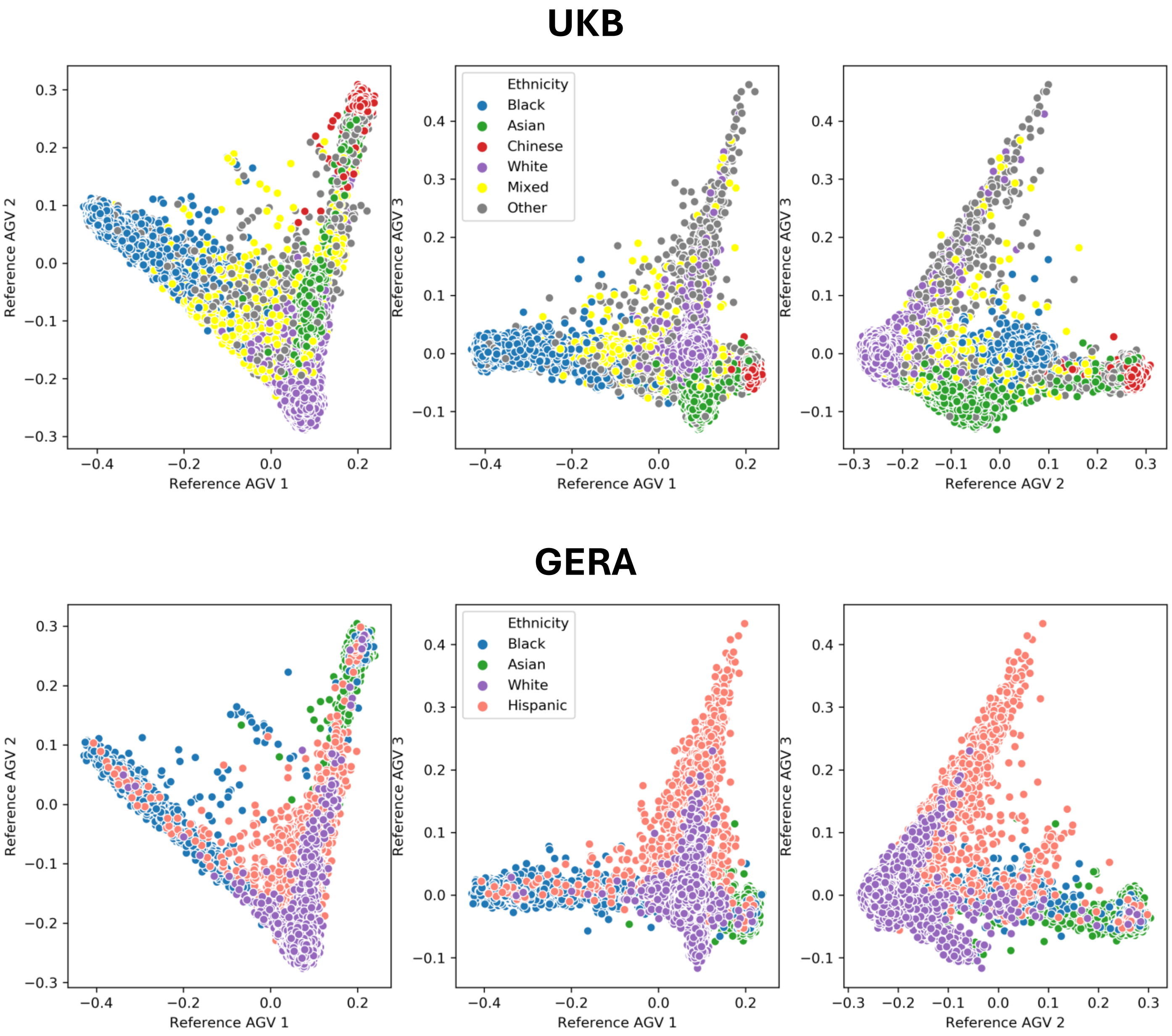


**Supplementary Figure 3. Projection of participants from the UK Biobank (UKB) and the the Resource for Genetic Epidemiology Research in Adult Health and Aging (GERA) onto the first three axes of genetic variation (AGVs) derived from 1KGP/HGDP (described in Figure 2).** Each point corresponds to a participant, coloured according to self-reported ethnicity. Projections for the four genotyping arrays in GERA have been combined.

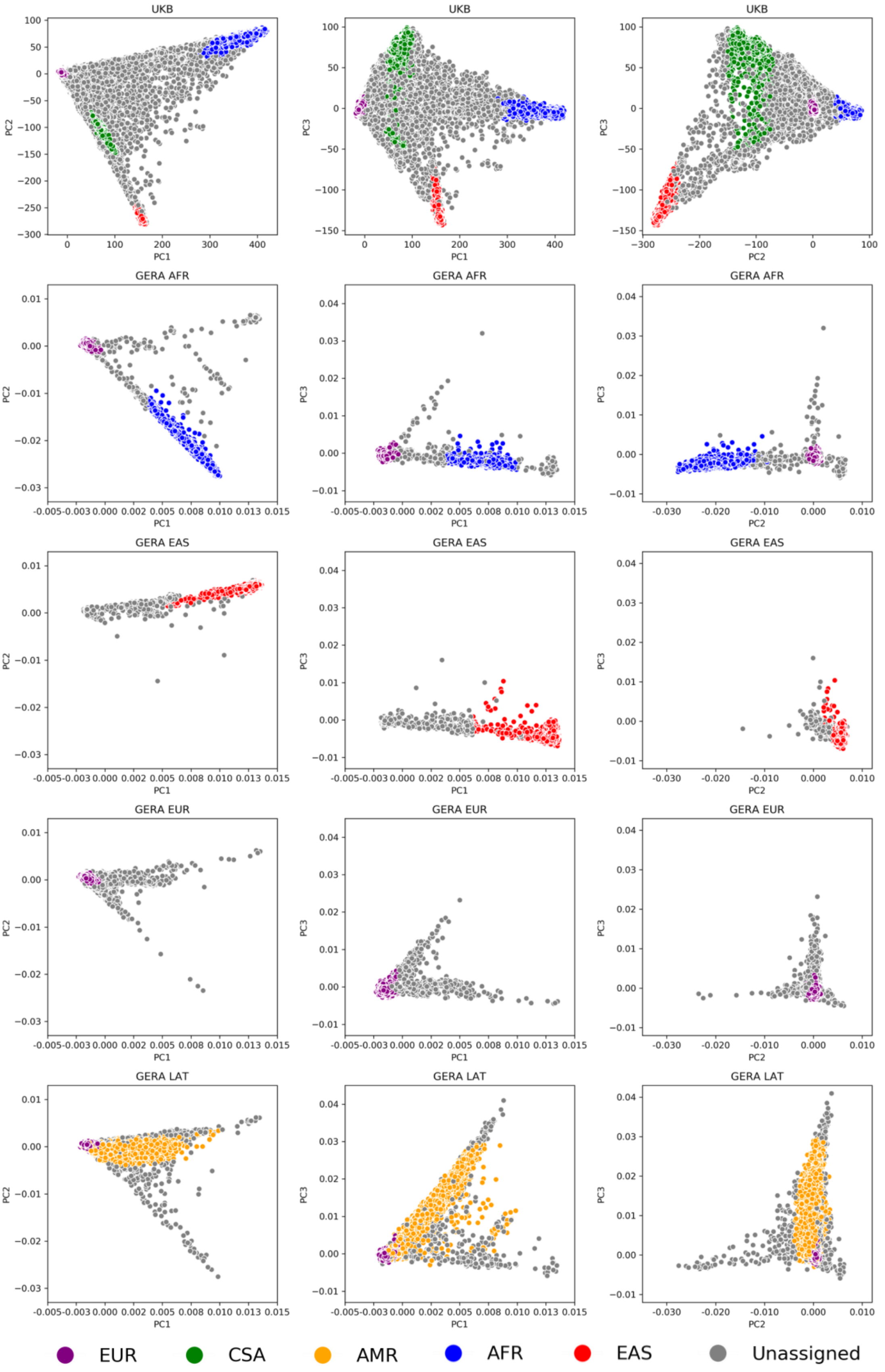


**Supplementary Figure 4. Assignment of participants to continental ancestry groups in ancestry-stratified GWAS analysis of T2D susceptibility in UK Biobank (UKB) and the Resource for Genetic Epidemiology Research in Adult Health and Aging (GERA, separately by genotyping array).** Each point corresponds to a participant, plotted according to their position on the first three genetic principal components, calculated separately for UKB and GERA (for each genotyping array). Participants assigned to a continental ancestry group are coloured: African (AFR), American (AMR), Central and South Asian (CSA), East Asian (EAS), and European (EUR). Participants not assigned to an ancestry group are coloured in grey.

### (a) Inclusive pooled analysis

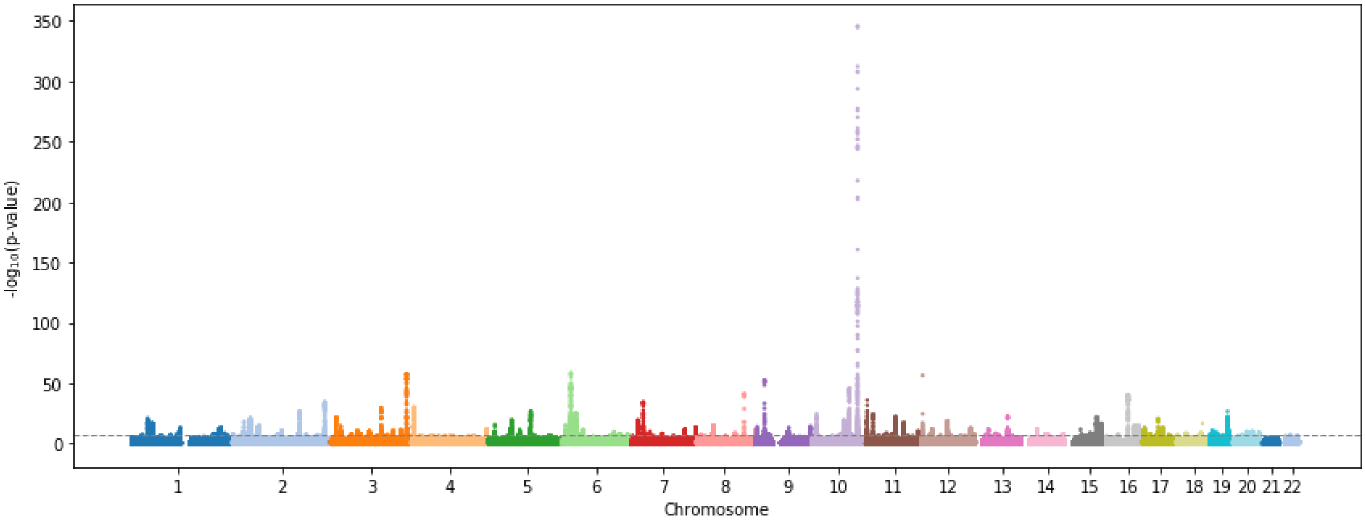


### (b) Ancestry-stratified analysis

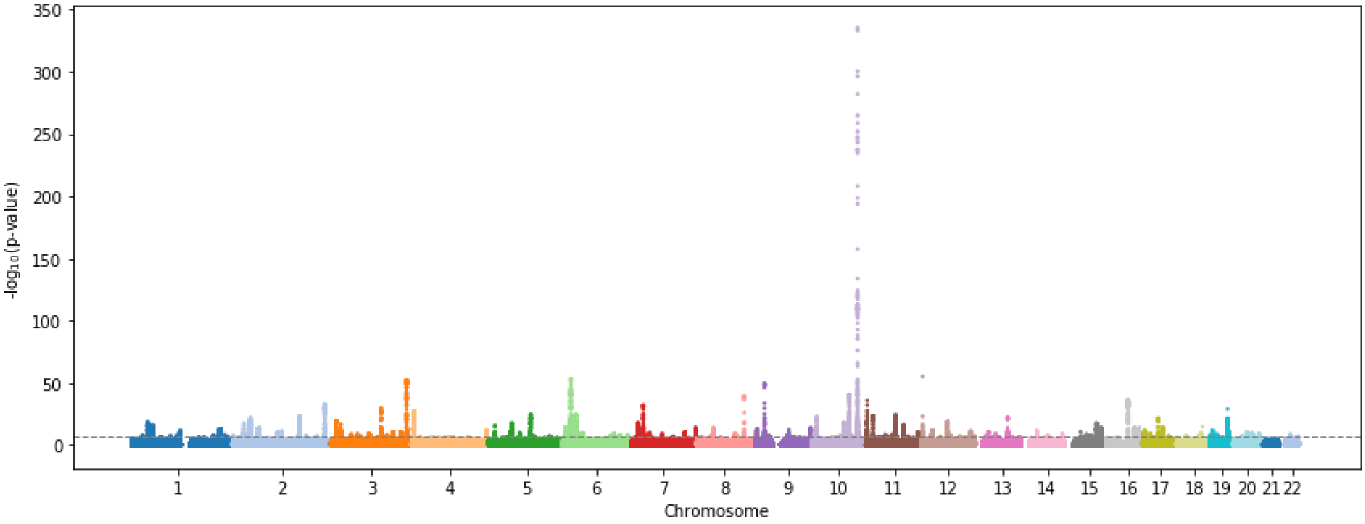


**Supplementary Figure 5. Manhattan plots of T2D association obtained from inclusive pooled analysis and ancestry-stratified analysis in the UK Biobank and the Resource for Genetic Epidemiology Research in Adult Health and Aging.** Each point represents a variant, plotted with their association *P*-value (on a -log10 scale) as a function of genomic position (NCBI build 37). Genome-wide significance ($P$<5x10$^{-8}$) is highlighted by the dashed horizontal line.

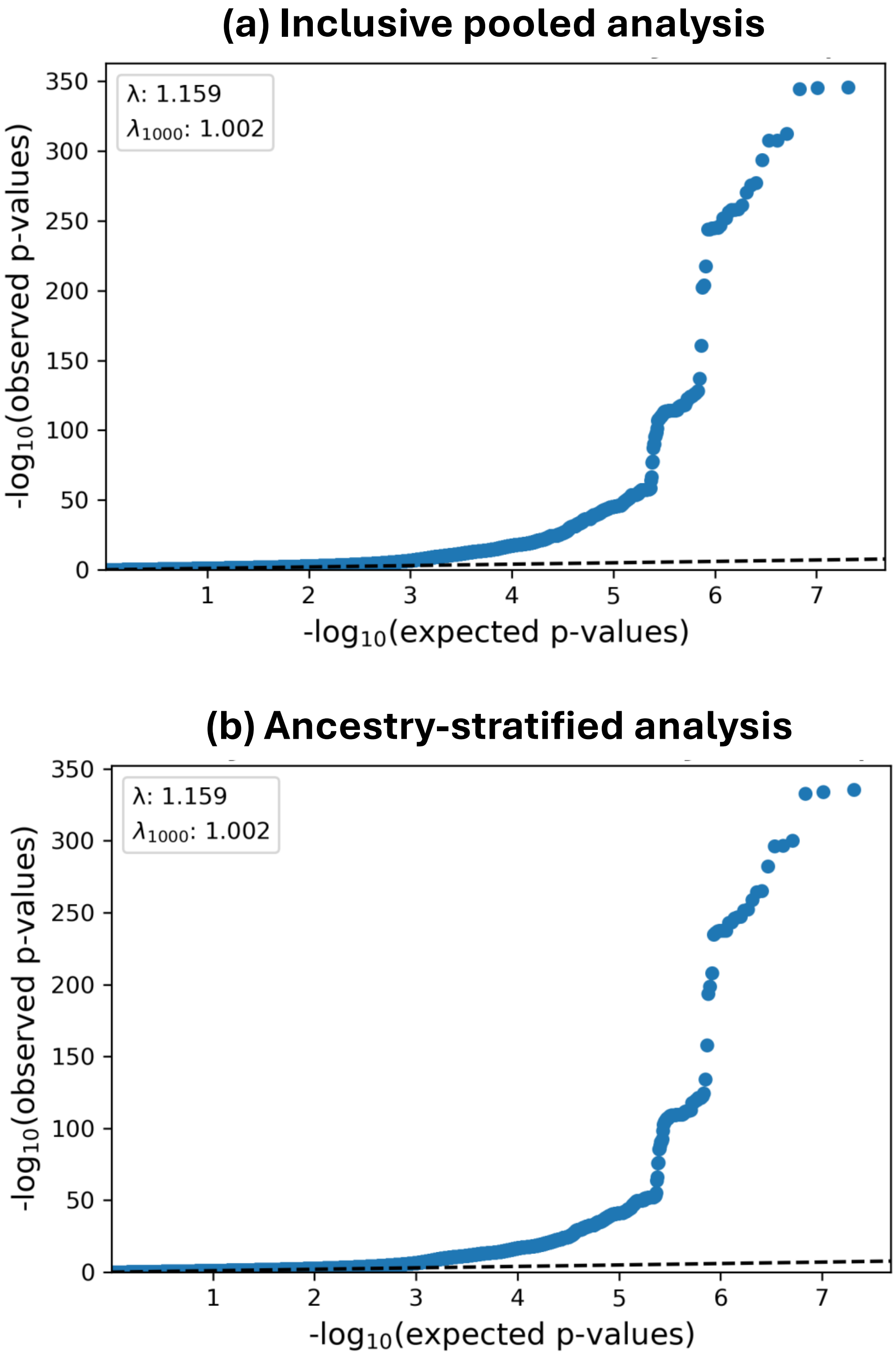


**Supplementary Figure 6. Quantile-quantile plots of T2D association obtained from inclusive pooled analysis and ancestry-stratified analysis in the UK Biobank and the Resource for Genetic Epidemiology Research in Adult Health and Aging.** Each point corresponds to a variant, plotted according to the observed P-value (on a $-\log_{10}$ scale) on the y-axis and the expected P-value (on a $-\log_{10}$ scale) under the null hypothesis of no association.

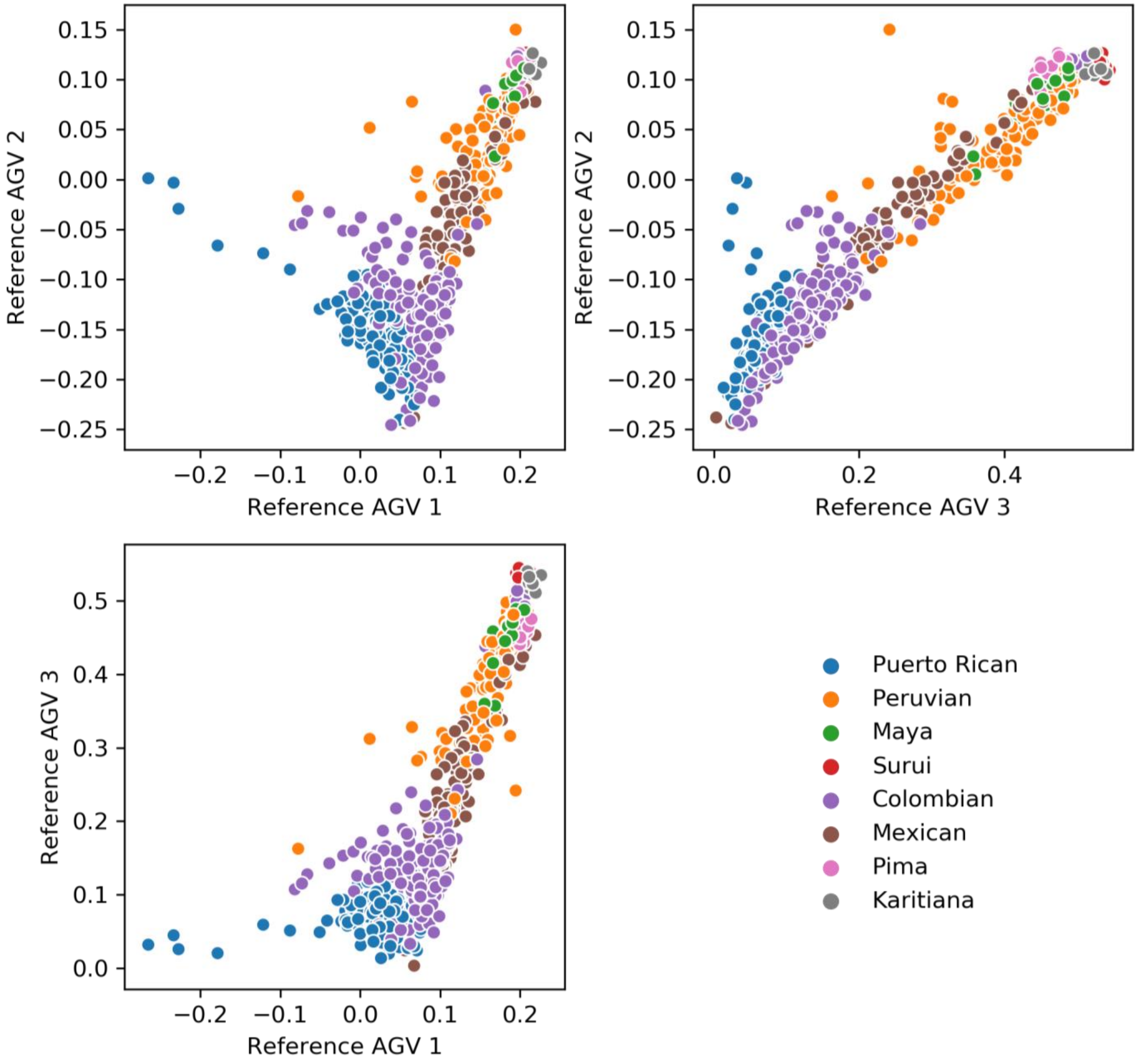


**Supplementary Figure 7. Placement of 1KGP/HGDP individuals from the American continental ancestry group on reference axes of genetic variation (AGVs).** In each panel, points correspond to individuals, plotted according to their position on the first three reference AGVs, obtained from principal components analysis of LD-pruned autosomal variants from whole-genome sequence data. Each individual is coloured according to self-reported population group. The first three AGVs emphasize a cline of genetic diversity for populations within the American continental ancestry group.

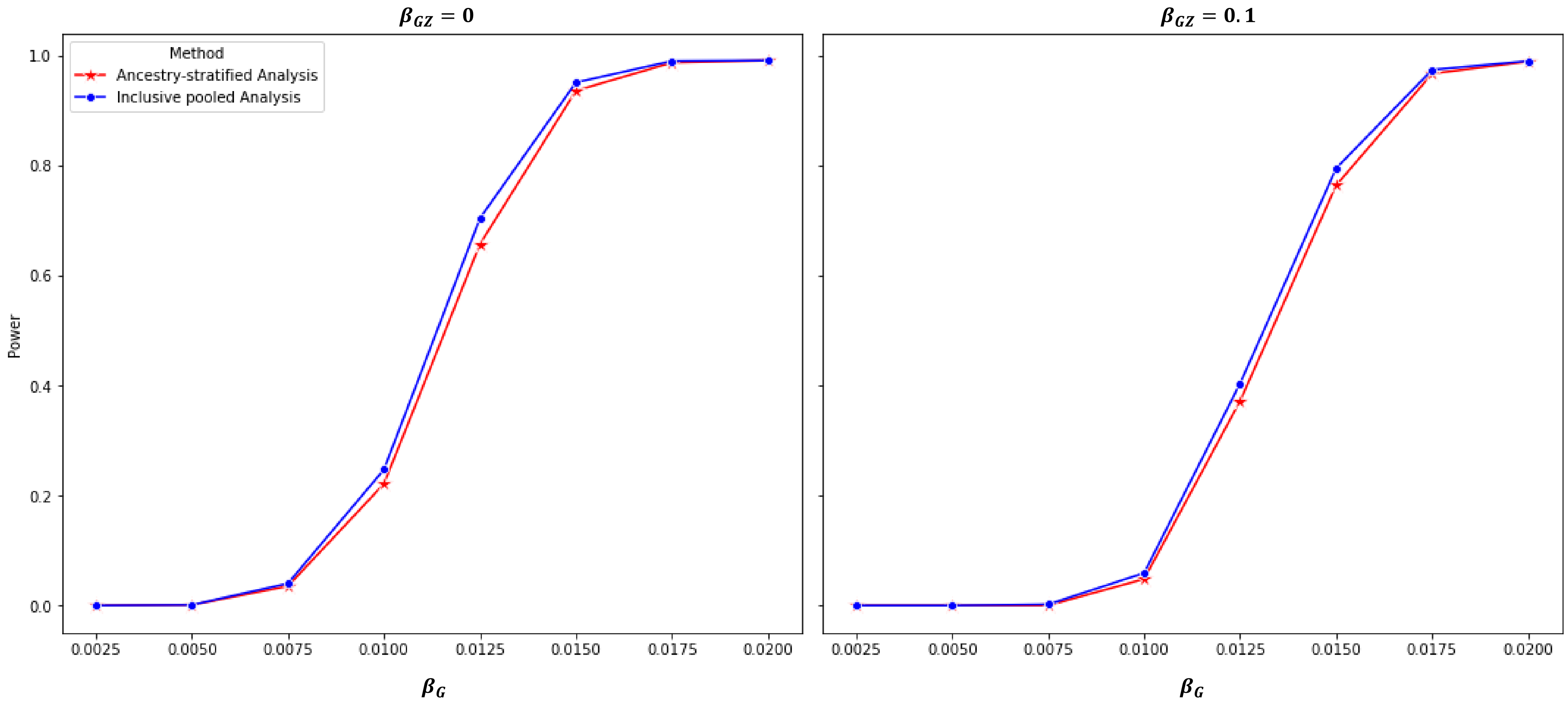


**Supplementary Figure 8. Power to detect association (at genome-wide significance, *P*<5x10$^{-8}$) with causal variants for inclusive pooled analysis and ancestry-stratified analysis based on participants in the UK Biobank.** Power is presented for each method as a function of the marginal effect of the causal variant, $\beta_G$, assuming a fixed marginal effect of an unobserved ancestry-correlated covariate, $\beta_Z = 0.1$, in the presence an absence of an interaction of the causal variant with the unobserved ancestry-correlated covariate, $\beta_{GZ} = 0.1$ and $\beta_{GZ} = 0$, respectively.

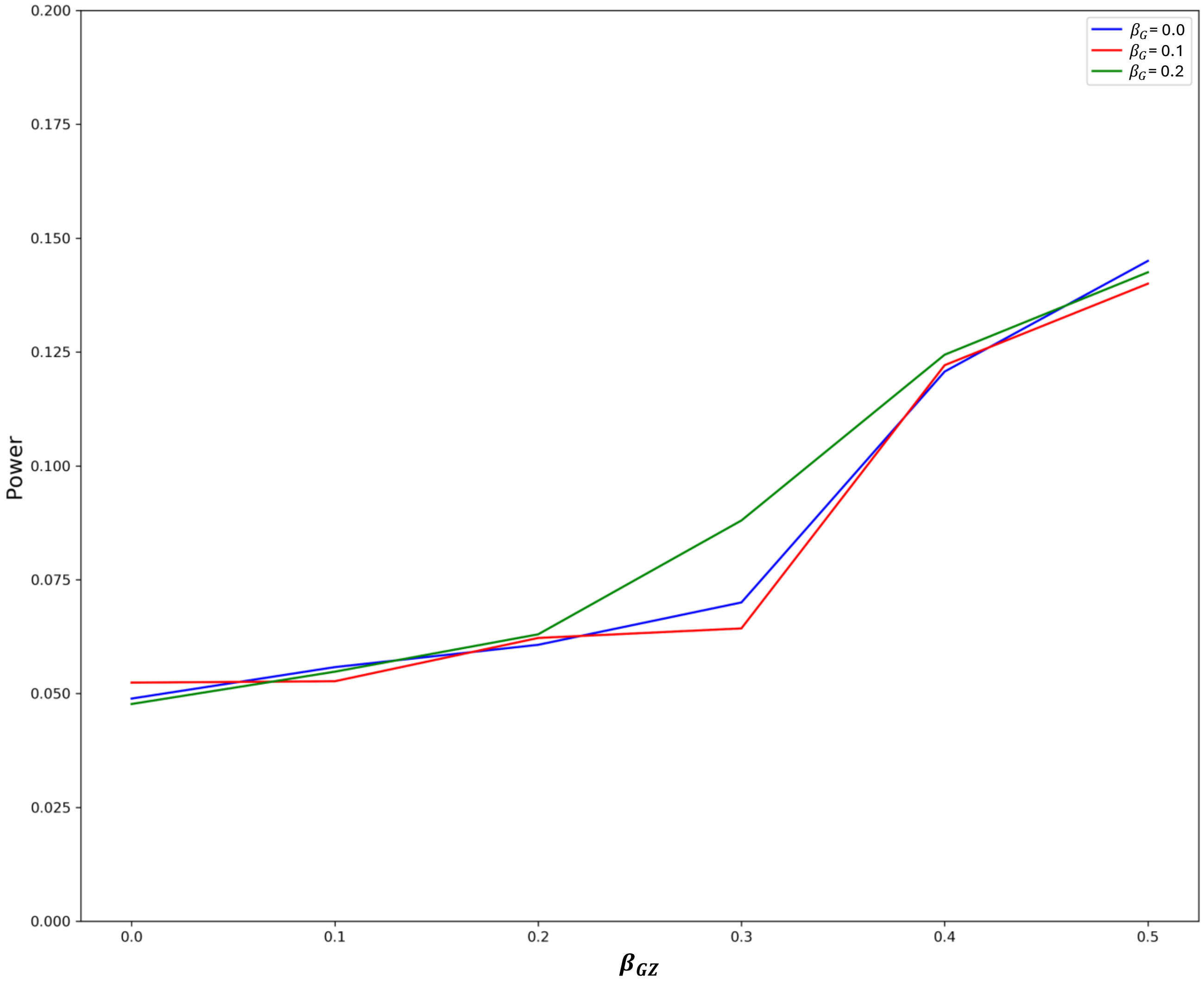


**Supplementary Figure 9. Power to detect ancestry-correlated heterogeneity (at nominal significance, *P*<0.05) at causal variants for inclusive pooled analysis based on participants in the UK Biobank.** Power is presented as a function of the interaction effect of the causal variant with an unobserved ancestry-correlated covariate, $\beta_{GZ}$, for four different marginal effects of the causal variant, $\beta_G = \{0, 0.1, 0.2\}$.
